# “At the end of the day I’d have to use a condom!”: A qualitative investigation of knowledge, motivations, and barriers to use of HIV pre-exposure prophylaxis among young gay and bisexual men in Nairobi, Kenya

**DOI:** 10.1101/2025.10.02.25336408

**Authors:** Carlos Cheruiyot, Peter Kaberia, Walter Nyagah, James Kang’ethe, Adrian D. Smith, Thesla Palanee-Phillips, Samuel Mwaniki

## Abstract

Despite availability of highly effective oral HIV pre-exposure prophylaxis (PrEP) for close to a decade, uptake and adherence among at-risk young men who have sex with men (YMSM) in Kenya, remain significantly low. This study investigated levels of PrEP knowledge, motivations, facilitators and barriers influencing PrEP initiation, persistence and adherence among YMSM, in efforts to optimise use and reduce risk of new HIV infections. In September 2021, 22 YMSM purposively selected from 248 YMSM who had previously participated in an integrated bio-behavioural sexual health survey, took part in semi-structured in-depth interviews. Interview guides were developed based on the Information-Motivation-Behavioural skills (IMB) model. Questions focused on each of the three IMB model constructs: information about PrEP, motivational factors influencing decision to use or not to use PrEP, and ability to initiate and adhere to PrEP. Data analysis generated three key themes: knowledge, motivation, and barriers to using oral and on-demand PrEP. Participants expressed its awareness and effectiveness in HIV prevention, but were concerned about PrEP- and HIV-related stigma, with the fear that family, friends, and potential romantic partners might perceive them as being promiscuous, or as living with HIV. Participants reported being capable of seeking PrEP services provided in MSM-friendly facilities, with public health facilities perceived as hostile considering the criminalization of, and societal stigma toward same-sex practices. These findings highlight the crucial need to re-strategize PrEP promotion not only as an effective but also a safe HIV-prevention option for at-risk populations. This study recommends expediting the integration of long acting lenacapavir and cabotegravir in the Kenyan health system as a means to diversify PrEP modalities and potentially reduce the stigma experienced in oral PrEP use.

## Introduction

The field of public health has achieved remarkable strides in reducing incidence rates of Human Immunodeficiency Virus (HIV), with the 2023 global data, showing a median HIV prevalence of 0.8% among adults aged 15 to 49 years. Nonetheless, gay, bisexual and other men who have sex with men (GBMSM - hereafter abbreviated as MSM) are disproportionately affected, with a global prevalence of 7.5% higher than the general adult population [1]. This disparity is evident in sub-Saharan Africa, where MSM have an HIV prevalence five times higher than men in the general population [2]. In Kenya adult MSM aged 15 to 49 have a reported prevalence of 25% [3] compared to 2.7% in the general male population of the same age group [4]. Young MSM (YMSM) in Kenya aged 18-24 years face distinct HIV-related vulnerabilities, with a prevalence of 3.6% - six times higher than that of young heterosexual men from the same age group [5]. A Nairobi-based 2021 study found that HIV diagnosis and care inequalities were largely age-drive, with young MSM and transgender persons (TP) aged 18 – 22 having median viral loads, 3.14 log higher than older peers (4.44 vs. 1.30 respectively, p = 0.0022), reflecting lower status awareness and care engagement [6]. The same study reported an HIV prevalence of 13% in this age group, suggesting that risk begins earlier in adolescence, making YMSM a priority group for targeted intervention.

HIV risk in Kenyan YMSM can be attributed to behavioural, structural, and biological factors. By the age of 18, most YMSM are likely to have completed their secondary education with a chance of joining higher-learning institutions [7], at which they have the freedom to interact with peers of similar sexual orientations [8], hence, the opportunity to engage in casual and other risky sexual behaviours such as sex work, condomless sex, group sex, and having multiple partners [5]. Homophobia and the criminalization of homosexuality in most sub-Saharan countries, including Kenya, facilitate punitive laws, stigma, and discrimination [9], which in turn, hinder MSM from seeking health services, including those geared toward HIV/STI prevention [10]. Biologically, the high probability of HIV transmission per act of receptive anal intercourse plays a central role in understanding the disproportionate HIV burden among MSM [11]. This is also the case with untreated bacterial sexually transmissible infections (STIs) among MSM, as they have the potential to enhance both HIV transmission and acquisition [12]. A recent study conducted in Kenya among YMSM showed that more than half of the study participants tested positive for at least one of five curable STIs (gonorrhoea, chlamydia, syphilis, M. *genitalium*, or T. *vaginalis*) [13], thus making YMSM in this setting a priority group for HIV prevention efforts.

In its commitment to reducing new HIV infections, Kenya, through the Kenya Pharmacy and Poisons Board, approved PrEP in 2015 [14]. Subsequently, the Ministry of Health incorporated both daily and on-demand oral PrEP in the guidelines on use of antiretroviral medicines for treating and preventing HIV infection in Kenya, followed by the introduction of PrEP in the public health system close to a decade ago [14,15]. Kenya recently approved two long-acting injectable PrEP options, cabotegravir (CAB-LA) (administered every two months) and lenacapavir (administered twice a year), although neither are widely available currently [18,19]. Since its launch, PrEP has been distributed free of cost across all counties in more than 2000 public and private health facilities with a cumulative initiation of 535,986 individuals as at 2024 from the initial 5,927 in 2017 [14,16]. This progress has relied heavily on external donor support, particularly from PEPFAR and USAID [17]. Proposed cuts to global health funding is already jeopardizing the availability of free PrEP and ARVs, especially for LGBTQ+-friendly NGOs that largely depend on donor funds.

Notably, despite the prospect of having these newer PrEP options, the currently available highly safe and effective daily oral or on demand PrEP option is still underutilized among YMSM. For instance, only 11.2% of eligible YMSM had ever taken PrEP, and only 5.8% were using PrEP as at the time of a recently conducted study in Kenya [13]. Whereas the minimal utilization of PrEP by young people in settings like South Africa has been attributed to factors such as lack of awareness and poor accessibility [20], there are insufficient studies to understand how YMSM perceive PrEP in the Kenyan context. This knowledge gap therefore necessitates the need for studies to explore YMSM’s attitudes and perspectives toward PrEP as an HIV prevention method. As part of this broader effort, this study aimed to qualitatively understand Kenyan YMSM’s knowledge, motivations and barriers to use of oral daily and on-demand PrEP. The insights gathered may in turn, provide helpful information to various stakeholders, including policymakers, service providers, and YMSM, in designing effective strategies focused on increasing PrEP acceptability, willingness to use, uptake, adherence and persistence among YMSM. Such measures may significantly contribute to reducing new HIV infections among YMSM, and contribute to the UNAIDS’ ambitious goal of ending AIDS as a public health threat by 2030.

## Materials and Methods

Study methods are detailed in the published study protocol [21], and summarized below:

### Ethics Statement

The study protocol was approved by University of the Witwatersrand Human Research Ethics Committee-Medical (Reference number. M200215) and the University of Nairobi-Kenyatta National Hospital Ethics and Research Committee (Reference number. P990/12/2019). The initial approval period was from March 5, 2020 to March 4, 2021, which was then renewed from March 5, 2021 to March 4, 2022. Prior to participation, participants were administered and completed written informed consent. Participants were given an amount of KES. 1,000 (approximately $10 at the time) to offset time spent during the study and travel-related expenses to and from the study site.

### Aim of **s**tudy

The study aimed to investigate YMSM’s knowledge, motivations and barriers to use of oral daily and on-demand PrEP as HIV prevention tools among higher education students in Nairobi, Kenya.

### Study design and approach

This was a qualitative study in which data were collected through in-depth interviews with YMSM. In-depth interviews were selected for their effectiveness in facilitating researchers to learn the experiences, opinions, and perceptions of study participants [22], providing a deeper understanding of the topic than would be possible through quantitative methods such as surveys [23]. This approach was also deemed more appropriate than focus group discussions (FGDs), given the sensitivity of the subject matter - sexual risk, which may be ill-suited to group settings due to potential discomfort or social desirability bias among participants. Data collection, analysis and interpretation was guided by the Information-Motivation-Behavioural Skills (IMB) model. The IMB model for PrEP posits that, if populations at heightened risk of acquiring HIV are knowledgeable about PrEP, motivated to act on their knowledge, and have the required behavioural skills to look for and initiate PrEP, then they can successfully overcome barriers to PrEP initiation and adherence [24].

### Study setting

This study was conducted in Nairobi, Kenya. As the capital city, Nairobi was suitable for accessing study participants as it offers an expansive urban landscape that hosts approximately 150 campuses of various universities and mid-level colleges [25], some of which are equipped with health facilities that serve the student population [26–28]. Nairobi is also home to approximately a number of non-governmental, community-based MSM-friendly clinics [29], alongside several community pharmacies [30] and public and private health facilities [31]: the collectively form a network through which YMSM may access a range of healthcare services, including PrEP.

### Study participants

Participants in this study (N = 22) were purposively selected from a larger sample of 248 students who had previously taken part in a quantitative respondent-driven sampling (RDS)- based integrated bio-behavioural survey, whose details are described elsewhere [5] [13] [32]. Tertiary education students were chosen as they were young, sexually active, and likely to have access to sexual health information. Study participants were ≥18; male sex at birth, in a tertiary institution in Nairobi, and had engaged in receptive or insertive oral and/or anal sex with an adult male in the past 12 months. In addition, only participants who tested negative for HIV during the RDS-based survey were considered eligible for this study. Fifty-two YMSM met the set eligibility criteria, from which 25 (∼ 10% of the 248 in the RDS-based survey) were purposively selected, with the number drawn from each RDS recruitment wave being proportional to the size of the wave. This sample size is based on previous studies being sufficient to yield thematic saturation. A similar method of choosing a tenth of participants from an RDS-based survey for subsequent interviews proved successful in a study conducted in Tanzania, even though it had a smaller sample of 10 participants [33]. Of the 25 invited YMSM for in-depth interviews, 22 participated, while three were unable due to personal reasons such as lack of time.

### Data collection process

Data were collected through semi-structured interviews designed to elicit discussion on each of the three constructs of the IMB model. The interview questions covered three main areas: YMSM knowledge of PrEP, their motivation for using/not using PrEP, and access PrEP-related services. Regarding YMSM’s knowledge about PrEP, questions posed to participants sought to find out what they knew about PrEP, and its target population e.g., “Tell me what you know about PrEP? Who do you think PrEP is for?” Under motivation, participants were prompted to answer questions on how they and other MSM in the community felt about PrEP and the possibility of taking a pill daily compared to on-demand PrEP and the prospected long-acting injectables (CAB and LEN) for HIV prevention: e.g., “How do you feel about PrEP? Would you take a pill daily to prevent getting infected with HIV?” Would you be interested to have an injection/shot with a drug to prevent HIV infection?" Interviewers further probed participants’ access to PrEP with questions such as; “Which health facility could you go to for PrEP? Where would you feel comfortable getting PrEP?”

The interviews were conducted in September 2021 in a private study office previously used to conduct the RDS-based survey, with each session lasting approximately 45 minutes. The sessions comprised of only the interviewer and a single participant in the room to ensure comfort and confidentiality. All interviews were recorded and transcribed verbatim, and in instances where participants responded in Swahili, translated into English to maintain accuracy and fidelity. Rigorous quality measures were applied, involving the random selection of transcripts by the interviewer for cross-referencing with the corresponding audio recording to confirm precision in transcription and translation.

### Data analysis

Data transcripts were organized and analysed using the NVivo software version 12, by members of the research team. Two team members independently identified recurring patterns in the data using an iterative coding process. Then, they compared the generated codes for agreement and engaged in a consensus-building process to decide whether to merge, eliminate, or introduce new codes, which further enhanced the validity and reliability of the data. The codes were then analysed thematically, with the resulting themes and sub-themes described and substantiated with illustrative interview excerpts. Excerpts were attributed to participants using codenames to maintain participant confidentiality.

### Trustworthiness of the data

The study employed four key criteria: credibility, dependability, transferability, and confirmability to ensure the data’s trustworthiness [34]. To establish credibility, rigorous criteria used in qualitative investigations were employed. Before collecting data, interviewers had built a strong rapport with study participants through previous interactions during the RDS-based survey. In addition, during data collection, an iterative approach was adopted, involving rephrasing previously asked questions to confirm and gain a deeper understanding of the information provided by participants. Member checking was conducted ‘on the spot’ by paraphrasing and summarizing participant responses at intervals and at the end of each topic of discussion. The study processes were thoroughly documented for dependability to provide adequate details, enabling future researchers to replicate the study. In ensuring transferability, the study methodology was described in sufficient detail to aid comprehension and facilitate the comparison of findings with similar studies. Finally, confirmability was ensured by supporting the findings with verbatim quotes from study participants.

## Results

### Participants’ characteristics

All participants (N=22) were cisgender males who identified as either gay or bisexual. Participants’ ages ranged between 18 and 24, with the majority (64%) depending on their parents or guardians for financial support. All participants were either in college or university in Nairobi’s metropolitan area, with the majority (73%) residing in campus housing and nearby hostels. Table 1 presents a summary of participants’ sociodemographic characteristics.

**Table 1:** Sociodemographic characteristics of YMSM^§^ (N=22)

| Characteristic | Categories | Frequency (%) |
| --- | --- | --- |
| Age | 18 – 20 | 45.5 (10) |
|  | 21 – 24 | 54.5 (12) |
| Gender identity | Cisgender male | 100.0 (22) |
| Sexual orientation | Gay | 72.7 (16) |
|  | Bisexual | 27.3 (6) |
| Institution attended | University | 50.0 (11) |
|  | College | 50.0 (11) |
| Type of institution | Public | 59.1 (13) |
|  | Private | 40.9 (9) |
| Type of course taken | Science, technology, engineering and mathematics | 54.5 (12) |
|  | Arts, business and humanities | 45.5 (10) |
| Year of study | 1<sup>st</sup> and 2<sup>nd</sup> year | 59.1 (13) |
|  | 3<sup>rd</sup> and 4<sup>th</sup> year | 40.9 (9) |
| Residence | College housing / rented hostel | 72.7 (16) |
|  | With family | 27.3 (6) |
| Ever attended boarding school | Yes | 86.4 (19) |
|  | No | 13.6 (3) |
| Main source of financial support | Parents / guardians | 63.6 (14) |
|  | Employment | 36.4 (8) |
| Owns a smartphone | Yes | 95.5 (21) |
|  | No | 4.5 (1) |

<sup>s</sup>YMSM: young gay, bisexual and other men who have sex with men

### Findings from qualitative interviews

The following sections explore the main themes that were defined from analysis of the interviews with YMSM. The themes include: YMSM’s knowledge and use of PrEP, key motivators for use or non-use of PrEP, and barriers to using PrEP. As a reminder, excerpts from the interviews were attributed to participants using pseudonyms, and their age in years, to maintain participant confidentiality.

#### YMSM’s knowledge and use of PrEP

Participants demonstrated a general understanding of the oral daily PrEP and its role in HIV prevention.

> “PrEP is a drug you take… if you are sexually active, so that it may prevent you from contracting HIV.” (005, YMSM)

> “PrEP should be used consistently for those people who are at a higher risk of getting HIV. That is sex workers, MSMs, and maybe any other person who might be having multiple partners.” (004, YMSM)

However, at the time of this study, only two participants were on PrEP, with most never having used, and others having started but discontinuing PrEP due to fatigue.

> “I have never used PrEP… but I know it is a good thing.” (065, YMSM) “I was using it but for now, I have stopped for a while.” (046, YMSM)

> “PrEP is a good thing. I used to be on PrEP but personally I just got tired.” (011, YMSM)

On the other hand, participants had limited awareness of on-demand PrEP. However, once informed about it, most felt that it would be easier to use, while others voiced that it would only work for planned sex.

> “It sounds very cool because if I have to take this and do away with my every day pill, I think this is the best way to go.” (069, YMSM)

> “At least with the daily pill, you’re covered because you never know where the risk of contracting HIV might come from. So, while on-demand PrEP isn’t bad, it’s not cool for me. I normally don’t plan for sex… like it’s random for me” (231, YMSM)

Similar observations were made when participants were introduced to the idea of taking long-acting injectable PrEP, with most expressing a preference to this option as it could potentially address the many challenges associated with the oral PrEP.

> “The injection is better. Because one can forget to take pills, so if you get an injection, you have full time protection. So even if you forget, you are just okay.” (025, YMSM)

> “I would prefer the injection… Because with that, I wouldn’t need to carry the pills wherever I go.” (120, YMSM)

#### Key motivators for use or non-use of PrEP

Participants highlighted several motivators for initiating PrEP, with the perceived high risk of contracting HIV being a common concern.

> “After going to the health clinic, I had exposed myself, and luckily my status was good, and I was told by the service provider that if I am exposed too much, I can be on PrEP so as to reduce the risk of getting HIV.” (231, YMSM)

Similarly, another participant explained how his friend’s experiences with those close to him contracting HIV influenced his decision to use PrEP:

> “He [a friend] told me about some of his friends he knew who got HIV in like less than six months of knowing them… he felt like HIV was almost everywhere… and that is why he was on PrEP to protect himself.” (015, YMSM)

Participants also felt that PrEP was an important prevention tool for individuals who prefer casual condomless sex, and those engaging in sex work.

> "My friends who use it, they are sex workers… I can say, it helps them and it is good they are using it" (172, YMSM)

> "PrEP has helped such people whereby if you don’t want to use condoms… those who don’t feel like using a condom but feel (unsafe), this is the best place I think they could land on." (069, YMSM)

> "Yeah. PrEP is a good thing for people who engage in risky behaviour. It just gives you the confidence that the risk of getting HIV is very low." (231, YMSM)

Mistrust in relationships was also mentioned as a reason for using PrEP due to the tendency of MSM having multiple sex partners, regardless of their relationship status.

> "People have very complicated relationships. You might be dating someone now, and saying you are a couple, both of you seem happy, but on the side, you also have other people that they don’t know of." (084, YMSM)

> “I’m sure most of us are using it, because mostly in MSM to get someone who has one partner is hard.” (046, YMSM)

Participants further mentioned that PrEP allowed for sexual relationships between HIV sero-different individuals and also reduced the risk of acquiring HIV from ill-meaning individuals who may want to knowingly spread HIV to their sex partners.

> “You may get a boyfriend who has HIV and you don’t, so the best way of doing it is for you to take it.” (025, YMSM)

> “Let’s say someone is HIV positive and maybe he doesn’t want to die alone so he wants to spread it… You know these people with bad intentions. So, taking PrEP can really help you just in case you end up having (sex with) one of those people.” (042, YMSM)

While most participants indicated being generally open to taking PrEP, many felt that they did not need it. A reason that stood out among participants was low perceived HIV risk as they either consistently used condoms, were faithful to one partner, or were not sexually active at the time of the study.

> "I’m not at that high risk like I can control it with… a condom…Yeah, so I feel like PrEP would… I don’t know…taking them daily is not ideal for me." (024, YMSM)

> "Yeah. I used it, a while back. But as of now, I am not using it. Mostly, nowadays, I stick to one partner and I use condoms." (085, YMSM)

> "I would use it if I were sexually active, but since I am not, I am not using it… because you can’t tell me that I will be taking PrEP every day and I am doing nothing." (084, YMSM)

Participant 084, argued against the need to use PrEP as it only prevented HIV, yet there were other STIs that needed to be prevented.

> "I mean I do not see it being of any use unless otherwise. There is the risk of STIs. That’s why I’d say I am not on PrEP because at the end of the day I’d have to use a condom. I mean if PrEP was like it prevents you from all the STIs and HIV as well, that could be perfect, but the fact that I’d have to use a condom even if I’m on PrEP, no!" (084, YMSM)

Participant 011, attributed his reluctance to taking PrEP to negative health impacts believed to be related to the drug such as weakening the immune system and the body in general.

> "This is my opinion… PrEP is a drug and it is something that you take every day and I think it weakens your immune system the same way… HIV patients… I was told like for HIV patients, there is a time their doctor tells them to stop taking (ARVs) for some time, I don’t know if that’s true… maybe it is just a rumour." (011, YMSM)

#### Barriers to using PrEP

One of the key barriers highlighted by participants was the stigma surrounding PrEP, which limits its uptake. Participants expressed concerns that family and friends might mistakenly assume they were living with HIV because PrEP pills resemble ARVs.

> "You can’t take the PrEP when you are in a place where people don’t know what it is. They will just think it is ARVs since it is in a container that looks like an ARV one… and the pills make the same noise in the bottles. Also, because you take it every day, they will think that you have HIV." (231, YMSM)

> "There’s also that shame… that stigma that comes with taking the pills every day because they are similar with ARVs… Yeah, and they (family / peers) see you taking it on a daily basis just like the way someone who has HIV takes them." (011, YMSM)

PrEP was also perceived as encouraging sexual disinhibition. Since PrEP is perceived to be highly effect, perceptions of it supporting people engaging with multiple sexual partners and risky behaviours, as some believed that users no longer had to worry about contracting HIV. This form of stigma was especially common within the YMSM community.

> "I would say he (friend) likes to explore. He takes PrEP… meets people and they just have sex. He is that type of a person who can’t settle. He just gets anyone he finds on the way." (057, YMSM)

> "I would also be concerned if my MSM friends get to know I am taking PrEP… Because they can also ruin your name." (042, YMSM)

> "They are actually judgmental… they are people you expect to be okay with it… I hear some people think you are a whore for using PrEP. They are like, why would you take PrEP if you don’t have a lot of sexual partners? Many people say it’s like a key to sleeping with different people, which is not the case." (015, YMSM)

Furthermore, difficulty negotiating PrEP use with current and potential sexual partners was a concern. Some feared that suggesting PrEP would make their partners question their fidelity, while others feared rejection from potential partners who associated PrEP use with high HIV risk.

> "I am faithful to this person but I don’t trust my partner… So, then again, I am afraid to, to tell him (about using PrEP) because he will say I’m doubting him or like I don’t trust him." (005, YMSM)

> "Student MSM, most of them they do know that there is PrEP but nowadays I think it’s very difficult for someone to find you with PrEP and someone to still trust you. You see like once you find PrEP you will be like, these things are for those who are at high risk so they’ll be like no, I’m not having sex with you." (024, YMSM)

Side effects from PrEP medicines were also highlighted as an obstacle to using PrEP. Some participants reported discontinuing PrEP due to persistent side effects such as dizziness and nausea.

> “People get their side effects differently… So, someone will vomit on the first day, vomit on the second day, vomit on the third day. Can you tell him to continue with the medication? He’ll tell you, "No! I do not want it" (003, YMSM)

> “The reason is that it normally caused me some dizziness, so I was kinda lazy every day. So, I stopped” (085, YMSM)

Participants also reported facing poor reception, discrimination, and confidentiality breaches at public health facilities, making it difficult to access PrEP services without fear of stigma.

> “There (public facilities), they don’t know that I am gay, so basically, they will ask me what is this for if I ask about PrEP, and stuff like that. If you disclose information that piques their interest, probability of you being exposed will be high.” (084, YMSM)

> "Those people in public hospitals don’t understand you, and the moment you mention anything about MSM, you don’t know how they will react. You fear they may treat you badly. Or even expose you in the community." (120, YMSM)

> “There’s PrEP yes, but they just feel like, why are you taking this, can’t you just use condoms… You might see me as safe, but how sure are you whatever I am telling you is true? Why do you want to ask me to go use a condom instead of giving me PrEP because I came for PrEP?” (003, YMSM)

As a result, participants preferred MSM-friendly clinics where they felt safe with friendly providers who also followed them up after initial visits to the facilities. However, participants noted that, unlike public facilities, MSM-friendly clinics were fewer and could be inaccessible if one travelled out of town.

> “I would just go to these non-governmental organizations or healthcare centres for the MSMs. Because I am free and feel safe when I go for my medications and healthcare there. Then the reception of those guys at the organizations is good. The clinicians are friendly and they follow up.” (120, YMSM)

> “If travelling, maybe they have gone to a place like Mombasa (Kenyan coastal city approximately 300 miles / 500km from Nairobi) and they didn’t carry enough PrEP, acquiring PrEP in other places is difficult since you can’t go to the public facility to be given PrEP. It means they have to stop for a while until they come to where (the MSM-friendly clinics in Nairobi) they normally get their PrEP.” (231, YMSM)

## Discussion

Our study aimed to explore YMSM’s knowledge, motivations and barriers to using PrEP for HIV prevention. Participants demonstrated a general understanding of PrEP and its role in HIV prevention; however, most were only familiar with the daily dosage and had little to no awareness of the on-demand and long-acting regimens. The primary reasons for using PrEP included perceived high HIV prevalence within the MSM community, lack of trust between sexual partners, preference for condomless sex, and engaging in sex work. Conversely, reasons for not using PrEP included a perceived low risk of HIV among participants, concerns about other STIs – demanding dual prevention efforts such as using barrier methods or condoms in addition to PrEP use, and misconceptions about PrEP. Notably, awareness and motivation alone seemed insufficient to facilitate PrEP use, as interested YMSM encountered significant barriers to access and use, including poor reception in public health facilities, HIV- and PrEP-related stigma, and medication side effects.

Participants demonstrated having access to adequate objective information regarding PrEP such as its availability, efficacy, dosage, and safety. However, this was not sufficient to influence PrEP uptake as most participants reported either never having used the daily PrEP regimen or having discontinued its use. According to Dubov et al.’s, IMB model for PrEP uptake, successful PrEP outreach is not only driven by the provision of objective information but also existing subjective information such as HIV prevention heuristics, the impact of PrEP on one’s sex life, and PrEP misinformation [24]. A reason that stood out for not using PrEP among study participants was the continued necessity of condom use. This finding suggests that YMSM view PrEP and condoms as mutually exclusive rather than complementary [35]. Similar sentiments have been observed in the past where a study in Kenya examining the "condom quandary" among PrEP users—though conducted in the general population, including heterosexuals in serodiscordant partnerships—found that participants questioned the emphasis on condom use given PrEP’s ability to reduce HIV transmission. A qualitative study among MSM in West Africa, further aligns with these findings, where participants failed to see the added value of PrEP over condoms [36]. These assertions, however, overlook the risk compensation effect, in which reduced or no condom use potentially increases vulnerability to other STIs [37].

Most participants demonstrated limited knowledge about on-demand or event-driven PrEP, despite its availability in the Kenyan health sector and its comparable effectiveness to daily PrEP [38]. Notably, once informed about on-demand PrEP, most participants preferred it over the daily regimen. This preference was also observed in a 2023 multi-country study conducted in South Africa, Uganda, and Zimbabwe, reporting high demand for on-demand PrEP among young men [39]. However, despite its advantages, on-demand PrEP is only effective for planned sexual encounters, as the first dose must be taken 24 hours before sex [38], whereas the daily regimen provides continuous protection for both spontaneous and planned encounters. The observed gap in accurate PrEP-related awareness highlights the need for innovative approaches to personalised PrEP information, perhaps using digital platforms including dating and sex apps [10].

Despite the high HIV prevalence among YMSM, most participants perceived their personal risk of infection as low. This perception echoes findings observed in a study on PrEP uptake among MSM in New York City, which found that while 80% of 505 participants were eligible for PrEP based on their HIV risk profiles, 78% of those eligible did not perceive themselves to be at high enough risk to justify its use [40]. Similarly, a systematic review of MSM’s relationship with women in sub-Saharan Africa found that MSM perceived their greatest risk of HIV acquisition to be from sex with women [41]. These beliefs contradict systematic review evidence estimating per-act HIV transmission risk to be 35 times higher for receptive anal sex and 2.8 times higher for insertive anal sex, compared to vaginal sex [42]. This perception may be shaped by public discourse, which often highlights the excess burden of HIV among women, while giving little attention to the significantly higher HIV prevalence among key populations such MSM. Consequently, this low-risk perception may lead to missed opportunities for PrEP initiation, as PrEP motivation and uptake depend on perceived HIV risk outweighing the rewards associated with engaging in risky behaviours [24].

PrEP being similar to ARVs in colour, packaging and dosage was highlighted as a hindrance for not using PrEP, especially when living with family and friends, as one would be assumed to be treatment for HIV. Similar sentiments were observed in a study conducted in Malindi, Kenya, among MSM who reported not using PrEP for fear of HIV-related stigma within their social networks [43]. In the same study, closeted individuals were concerned about their family and friends finding out about their sexuality and sexual behaviour if found using PrEP. These reasons are not confined to the African context as a study conducted in Alabama, USA, found that Black YMSM attributed negative perceptions towards PrEP to homonegativity in their communities, since using PrEP would cause the unintentional consequence of outing individuals who are not comfortable with their sexual identity, especially if in relationships with women [35]. Interestingly, PrEP use was perceived negatively even within the MSM community as they too, associated PrEP with risky behaviours, thus judging each other for using PrEP.

Discrimination and criminalization of same-sex practices also proved to be a challenge preventing YMSM from accessing and using PrEP. For instance, our study participants were only comfortable seeking PrEP services from MSM-friendly facilities. This is mostly due to public facilities shying from providing services to individuals engaging in same-sex activities and MSM who prefer to keep their sexual activities private, as it is a criminal offense in Kenya and most sub-Saharan countries [9]. These findings amplify those from our previous work on YMSM’s engagement with healthcare, where participants reported stigma, discrimination, and prejudice in public and institution-based health facilities [10]. Also, while YMSM may enrol for PrEP in MSM-friendly clinics, these clinics are only within cities and established towns, hence not accessible to YMSM in rural settings, thus preventing potential PrEP users from accessing this service. Notably, while the Kenyan penal code criminalizes same-sex practices, the constitution guarantees the right to health for its citizens [44]. Therefore, healthcare providers are obligated to ensure that every individual, regardless of their sexuality, can access quality health care, including HIV prevention services. It is also imperative for healthcare providers in both public and institution-based health facilities to undergo training and sensitization on the unique health needs of YMSM, and how to provide culturally competent services to this population in a non-judgmental manner [45].

However, it is important to note that even with sufficient knowledge and motivation to use PrEP, some participants still struggled to navigate discussions about PrEP with healthcare providers, family, peers, and partners. They also faced challenges in adhering to the daily dosage and managing side effects, ultimately making it difficult to sustain PrEP use in the long run. This aligns with Dubov et al.’s assertion that being well-informed and highly motivated is not always enough as individuals may still face difficulties initiating and maintaining PrEP if they lack the necessary behavioural skills to do so [24]. These behavioural skills, also referred to as self-efficacy, refer to a person’s belief in their ability to control a behaviour and their confidence in making the choice to follow through with it [46]. Although this highlights the crucial role of the behavioural skills concept in the IMB model, since it directly influences whether knowledgeable and motivated YMSM can successfully initiate and adhere to PrEP, intentions do not always lead to action [47]. To bridge the gap between intention and sustained health behaviour, action planning and coping planning have been proposed as effective planning constructs [24]. Action planning involves specifying when, where, and how to take PrEP consistently, while coping planning prepares individuals for potential challenges, such as forgetting doses, dealing with side effects, or addressing concerns from partners [48,49]. Nevertheless, it is essential to recognize that the goal of PrEP programs is not universal initiation and adherence to a specific regimen. Rather, PrEP regimens should be viewed as HIV prevention options that should be matched to an individual’s preferences, needs, and perceived risk levels. From this standpoint, the “right” prevention method may vary between individuals and across time as circumstances shift. Thus, important to understand how men may move in and out of PrEP use based on changes in risk and options.

Compared to daily and on-demand-oral PrEP, long-acting injectables (CAB-LA and lenacapavir), were highly preferable to YMSM. As they have been shown to be more effective in preventing HIV infection among men [50], and potentially reduce pill-related stigma and adherence issues. CAB-LA was approved as a PrEP modality by the Kenya Pharmacy and Poisons Board in 2024, although implementation remains limited by high costs and low availability [18]. Notably, injectable lenacapavir, approved mid-2025, offers a more promising alternative. Administered subcutaneously every six months, lenacapavir has demonstrated superior efficacy compared to existing PrEP regimens, among men and gender diverse persons [51]. While we recommend the expedited implementation of CAB-LA and lenacapavir in Kenya to address challenges associated with daily or on-demand PrEP, we acknowledge challenges associated with this strategy. These include high costs compared to oral PrEP, with lenacapavir being projected to cost $35-46) per person-year [52], which is highly unaffordable to unemployed individuals and low-income earners, considering Kenya’s current minimum wage is currently at Ksh 8000 (61 dollars) [53]. The other challenge is integrating the injectable into existing health systems, particularly with respect to health care worker-training, and logistics related to refrigeration of the drugs. Moreover, the continued need for oral PrEP after the last injection also presents logistical and adherence issues [54].

This study had a limitation in that, it was conducted in Kenya’s capital city which hosts a variety of healthcare facilities, including MSM-friendly clinics that provide access to PrEP-related information and services to YMSM. As such, the findings may not represent YMSM in rural settings with no access to sexual health services from MSM-friendly facilities. We also acknowledge that the tertiary-level students who participated in this study are not representative of the broader YMSM population in Kenya. As such, issues related to misperceptions, misinformation, and self-efficacy in addressing sexual health needs are likely to be even more pronounced among less educated or less privileged individuals.

## Conclusions

It is recommended that YMSM receive targeted education on PrEP to facilitate informed decision-making, with digital platforms including dating and sex apps, leveraged to ensure widespread dissemination of accurate information. Additionally, PrEP promotion strategies should shift from positioning it as a tool for at-risk populations, to a general HIV prevention intervention, thereby reducing stigma, especially within an already stigmatized population. Healthcare providers in both public and institution-based health facilities should also undergo training and sensitization on the unique health needs of YMSM to provide culturally competent, non-judgmental services. Furthermore, we advocate for the expedited implementation of CAB-LA and lenacapavir within the Kenyan health system. These long-acting injectables would address the challenges associated with daily and on-demand dosing while simultaneously reducing adherence issues and stigma related to taking PrEP pills, enhancing convenience, acceptability, and scalability, given its bi-annual dosing schedule. Lastly, we recommend incorporating supplementary preventive measures such as doxycycline post-exposure prophylaxis (doxy-PEP) to mitigate the risk of bacterial STIs – particularly in instances of condomless sex. A comprehensive approach that integrates these interventions would significantly strengthen HIV prevention efforts while advancing sexual health equity for YMSM.

## Data Availability

Data cannot be shared publicly because they are qualitative interviews which contain personal and potentially identifiable information from members of both a criminalized and stigmatized population, and participants have consented to publication of anonymized quotes only. Data are available from the corresponding authors on reasonable request for researchers who meet the criteria for access to confidential data, as determined by the approving ethics committees i.e. University of the Witwatersrand Human Research Ethics Committee Medical (via) and University of Nairobi-Kenyatta National Hospital Ethics and Research Committee (via).

## Acknowledgments

The authors would like to thank Rhoda Wanjiru, Evelyn Ombunga, Elizabeth Rwenji and Ibrahim Lwingi for supporting the data acquisition process. We are grateful to Joyce Kafu and Jacqueline Akinyi for transcribing the audio-recordings, and Hillary Koros for guiding the data analysis process. Lastly, we thank all participants for making the study possible.

